# COVID-19 in Iran, a comprehensive investigation from exposure to treatment outcomes

**DOI:** 10.1101/2020.04.20.20072421

**Authors:** Mohammad Ali Ashraf, Nasim Shokouhi, Elham Shirali, Fateme Davari-tanha, Omeed Memar, Alireza Kamalipour, Ayein Azarnoush, Avin Mabadi, Adele Ossareh, Milad Sanginabadi, Talat Mokhtari Azad, Leila Aghaghazvini, Sara Ghaderkhani, Tahereh Poordast, Alieh Pourdast, Pershang Nazemi

## Abstract

**Purpose:** There is a growing need for information regarding the recent coronavirus disease of 2019 (Covid-19). We present a comprehensive report of Covid-19 patients in Iran.

**Methods:** One hundred hospitalized patients with Covid-19 were studied. Data on potential source of exposure, demographic, clinical, and paraclinical features, therapy outcome, and post-discharge follow-up were analyzed.

**Results:** The median age of the patients was 58 years, and the majority of the patients (72.7%) were above 50 years of age. Fever was present in 45.2% of the patients on admission. The most common clinical symptoms were shortness of breath (74%) and cough (68%). Most patients had elevated C-reactive protein (92.3%), elevated erythrocyte sedimentation rate (82.9%), lymphocytopenia (74.2 %) on admission. Lower lobes of the lung were most commonly involved, and ground-glass opacity (81.8%) was the most frequent finding in CT scans. The administration of hydroxychloroquine improved the clinical outcome of the patients. Lopinavir/ritonavir was efficacious at younger ages. Of the 70 discharged patients, 40% had symptom relapse, (8.6%) were readmitted to the hospital, and 3 patients (4.3%) died.

**Conclusion:** This report demonstrates a heterogeneous nature of clinical manifestations in patients affected with Covid-19. The most common presenting symptoms are non-specific, so attention should be made on broader testing, especially in age groups with the greatest risk and younger individuals who can serve as carriers of the disease. Hydroxychloroquine and lopinavir/ritonavir (in younger age group) can be potential treatment options. Finally, patients discharged from the hospital should be followed up because of potential symptom relapse.

## Introduction

Coronaviruses are the second cause of the common cold after rhinoviruses.^1^ Human coronavirus pathogens can cause a wide range of diseases from the common cold to severe pneumonia. Two previous large-scale pandemics of coronavirus infections in 2002-2003 [coronavirus-severe acute respiratory syndrome(SARS)] and 2012 [coronavirus-Middle East respiratory syndrome (MERS)] had severe global health impacts.^2,3^ The recent coronavirus disease of 2019 (COVID-19) has stricken the global health and the economy even more than the previous ones. It has spread to more than 209 countries/territories and has infected more than a million people around the world. Iran has been one of the most severely affected countries by the virus.^4,5^

Previous studies described the clinical and demographic characteristics of the disease. Information regarding the transmission pattern is mostly related to China. There is also, a lack of information about the potential treatment outcomes, and post-hospitalization follow-up in the literature.^6-9^

This study is one of the first reports of COVID-19 patients from Iran. We reported detailed information about the potential source of exposure, household contact information, outcomes of potential therapies, and post-discharge follow-up, as well as demographic, clinical, and paraclinical characteristics.

## Methods

### Patients and study overview

Medical records of suspected cases of COVID-19 from February 22, 2020, to March 5, 2020, admitted to the YAS hospital affiliated to Tehran University of Medical Sciences, were reviewed. Our hospital was the first center in Tehran to care for adult COVID-19 patients. A suspected case was defined as a flu-like syndrome / or symptomatic patient along with radiologic pulmonary findings. Data of patients for whom the results of reverse-transcriptase-polymerase-chain-reaction (RT-PCR) were not available was excluded from the study. COVID-19 was confirmed using RT-PCR of nasopharyngeal specimens. This study was approved by the Tehran University of Medical Sciences (TUMS) ethics committee (IR.TUMS.VCR.REC.1398.1036).

Informed consent was obtained from all patients or their first-degree relatives in unconscious patients.

## Data sources

Patients who came to the hospital were examined by an infectious-disease specialist, and classified into three groups according to disease severity based on Iran’s national guideline for the diagnosis and treatment of COVID-19 in outpatients and inpatients (Figure1).^10^ Patients assigned to moderate or severe infection group were admitted to the hospital. Patients’ occupation, travel history within the past 14 days, household contact information, demographic characteristics, potential source of exposure, influenza vaccination history, current list of medications, past medical history, social history, and the use of preventive measures were determined.

**Figure 1.**
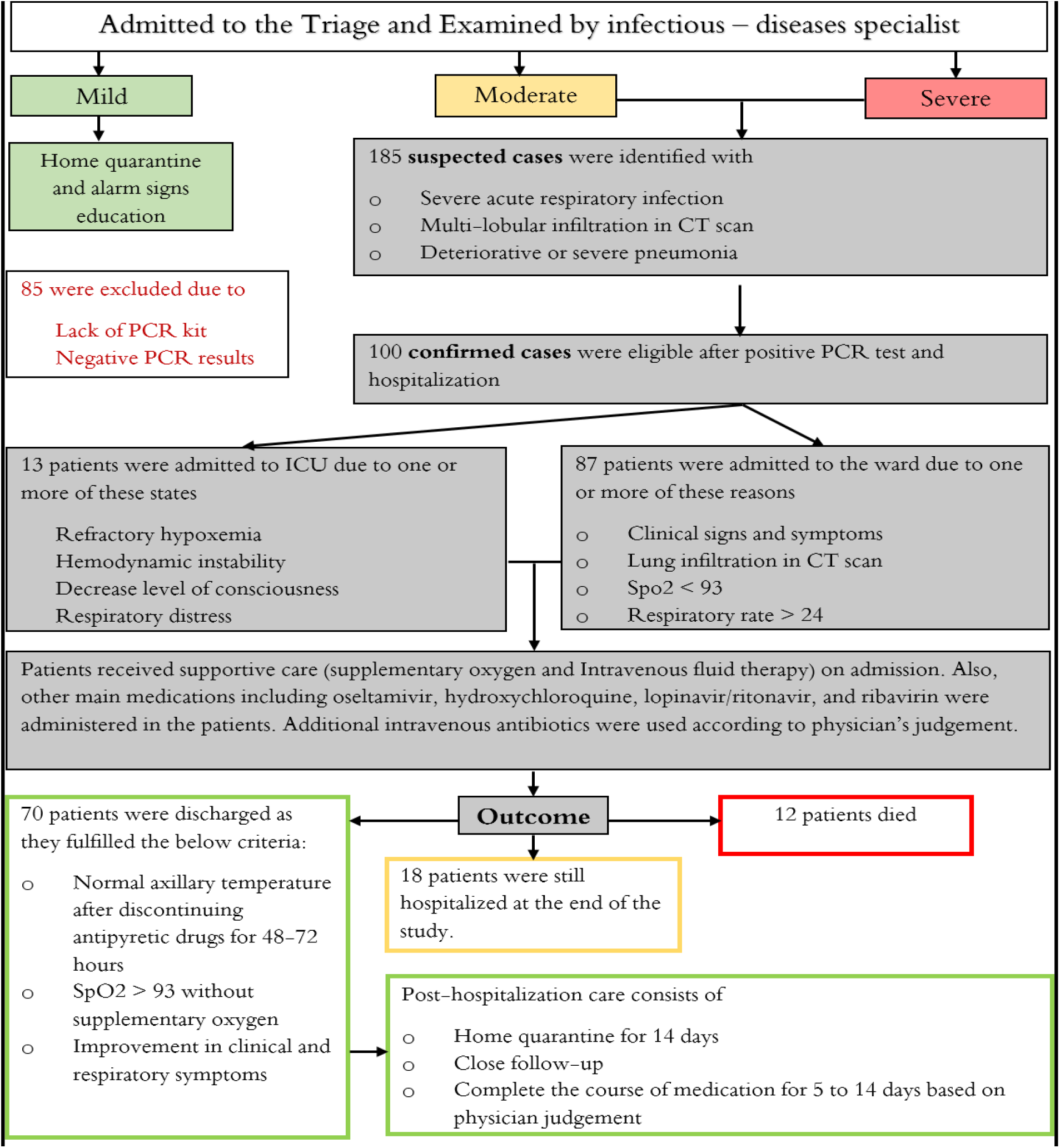
Flow Chart with an Overview of Study Steps. Patients were classified into three groups of mild, moderate, and severe disease. Treatment regimen and admission/discharge criteria were according to Iran’s national guideline for novel coronavirus infection. The definition of mild, moderate, and severe disease was as below according to the national guideline: Patients with a flu-like syndrome with/without fever, who did not have any signs of infiltration in lung imaging were classified as having mild disease. The moderate group was defined as symptomatic patients with pulmonary infiltration or at least one of the admission criteria, as explained in the figure. The severe group constituted patients who have at least one of the following criteria: 1. Reduced consciousness; 2. Respiratory rate (RR) ≥30; 3. Blood pressure (BP)BP<90/60; 4. Multi lobular infiltration; 5. Hypoxemia.

History of present illness and comprehensive review of systems were taken, and a complete physical examination was done. Clinical laboratory studies and chest computed tomography (CT scan) were requested on the first day of admission according to infectious disease specialist recommendations. We collected hospitalization data using patients’ paper medical records.

Available CT scans were reported by a radiologist and scored for severity and location of involvement. The final reports were reviewed by an infectious disease specialist and a pulmonologist.

The main treatment medications included oseltamivir (75mg twice daily), hydroxychloroquine (200mg twice daily / 400 mg single dose when combined administration with Lopinavir-Ritonavir), Lopinavir-Ritonavir (400 mg Lopinavir-100 mg Ritonavir twice daily), and Ribavirin (1200 mg twice daily) according to the national guideline.^10^

Also, we conducted a telephone survey of patients who were discharged from the hospital. A questionnaire was developed to follow patients for 14 days post-discharge. We asked patients about the episodes of symptom relapse, the need for hospital readmission, and whether they completed 14 days of home quarantine after discharge. Discharged patients were followed up to March 19, 2020.

### Study outcomes

The critical situation of the patients, which was defined as admission to an intensive care unit, the use of mechanical ventilation, or death, was considered as a primary composite endpoint. We compared demographic characteristics, hospitalization data, and potential treatment outcomes in critically ill and non-critically ill patients. Post-discharge follow-up was reported from the discharged patients.

### Study definitions

Index patient was defined as the first person in a household diagnosed with Covid19 using RT-PCR. The incubation period was calculated from the time between the last potential exposure and the time showing the first disease symptoms.

Lung lobar scores were calculated using a scoring system giving each five lobes a score graded from 0-4 according to the severity of the involved lobe. (0 = not involved; 1= up to 25 % involvement; 2 = 26-50 % involvement; 3 = 51-75 % involvement; 4 = 76-100 % involvement). The sum of all lobar scores combined is defined as the total lung score, which estimates the severity of the entire lung involvement (provides a score between 0-20). Lower lobes score was defined as the sums of right lower lobe and left lower lobe scores (provides a score between 0-8). The middle lobe score was defined as the right middle lobe score (provides a score between 0-4). Upper lobes score was defined as the sums of right upper lobe and left upper lobe scores (provides a score between 0-8).

### Laboratory confirmation

Laboratory confirmation of SARS-CoV2 was performed in the National Influenza Center located at the School of Public Health, Tehran University of Medical Sciences. Nasopharyngeal swab specimens were collected from hospitalized patients using Dacron sterile swabs and placed in 2 cc viral transport media and sent to the laboratory in cold condition. All samples were subjected to RNA extraction with High Pure Viral Nucleic Acid Kit (Roche, Germany) according to the manufacturer’s instructions. Real-Time (RT)-PCR was used to detect the presence of SARS-CoV2 with kits (ModularDx Kit, Wuhan CoV E & RdRP genes) provided by WHO targeting the E region for screening and RNA dependent RNA polymerase for confirmation. Invitrogen SuperScript III One-Step RT-PCR System with Platinum Taq DNA Polymerase was used for PCR. For each reaction, 12.5 µl reaction mix, 1 µl RT enzyme, 0.5 µl primer, probe mix and 5.6 µl PCR grade water was added to 5 µl RNA template. Cycling conditions for amplification of E and RdRP genes were 50 °C for 30 min, 95 °C for 2 min then 45 cycles of 95 °C for 15sec and 58°C for 30sec. A cycle threshold value of less than 36 Ct was defined as a positive test result.^11^

### Statistical analysis

Non-parametric tests (including Fisher’s exact test, Mann-Whitney U test, and Friedman test) were used to analyze data. Cross tabulation and Fisher’s exact test were used to investigate the relation between the binary variables. Mann-Whitney U test was applied to compare the quantitative variables between the two groups, and the median and interquartile range (IQR) were presented with the results. In the CT scan analysis, the Friedman test was used to compare between different lung lobes involvement and comparison of triple accumulative scores. In addition, logistic regression was used to estimate the effect of the treatment on an odds ratio (OR) scale using the backward Wald elimination of variables. In the regression model, the response variable was considered as a binary variable with either 0 or 1 (1 in case of discharge and recovery, and 0 in case of death). All of the administered medications (hydroxychloroquine, lopinavir/ritonavir, ribavirin, and antibiotics were entered into the regression model as binary and independent variables. Patients’ age and coexisting disorders (including hypertension, diabetes, and COPD/asthma) were considered as covariate variables, and the interaction between age and patient’s condition (critically ill vs. non-critically ill), and medications (hydroxychloroquine, lopinavir/ritonavir, and ribavirin) were included in the model. Also, in order to examine the simultaneous effect of hydroxychloroquine and azithromycin, the interaction of these two variables was considered in the model. All analysis was performed using SPSS software, version 23 (IBM).

## Results

In this study, we included 100 hospitalized patients out of 185 admitted patients from February 22, 2020, to March 5, 2020. Figure 2 shows the distribution of the index patients in 22 districts of Tehran and the surrounding areas/cities. District 2 was the most affected district in Tehran, followed by district 12,5,8, and 3. Findings show that 37% of the patients either lived in or visited these neighboring areas within the 14 days prior to admission. Five of these patients were linked to the city of Qom, the epicenter of the disease in Iran.^12^ Recent potential exposures, household contact information, demographics, clinical characteristics, laboratory, and radiologic findings, and patients’ outcomes were extracted as shown in table1. Clinical, paraclinical, and treatment outcome was also compared in two groups of discharged and deceased patients (see Table S1 in the Supplementary Appendix).

**Figure 2.**
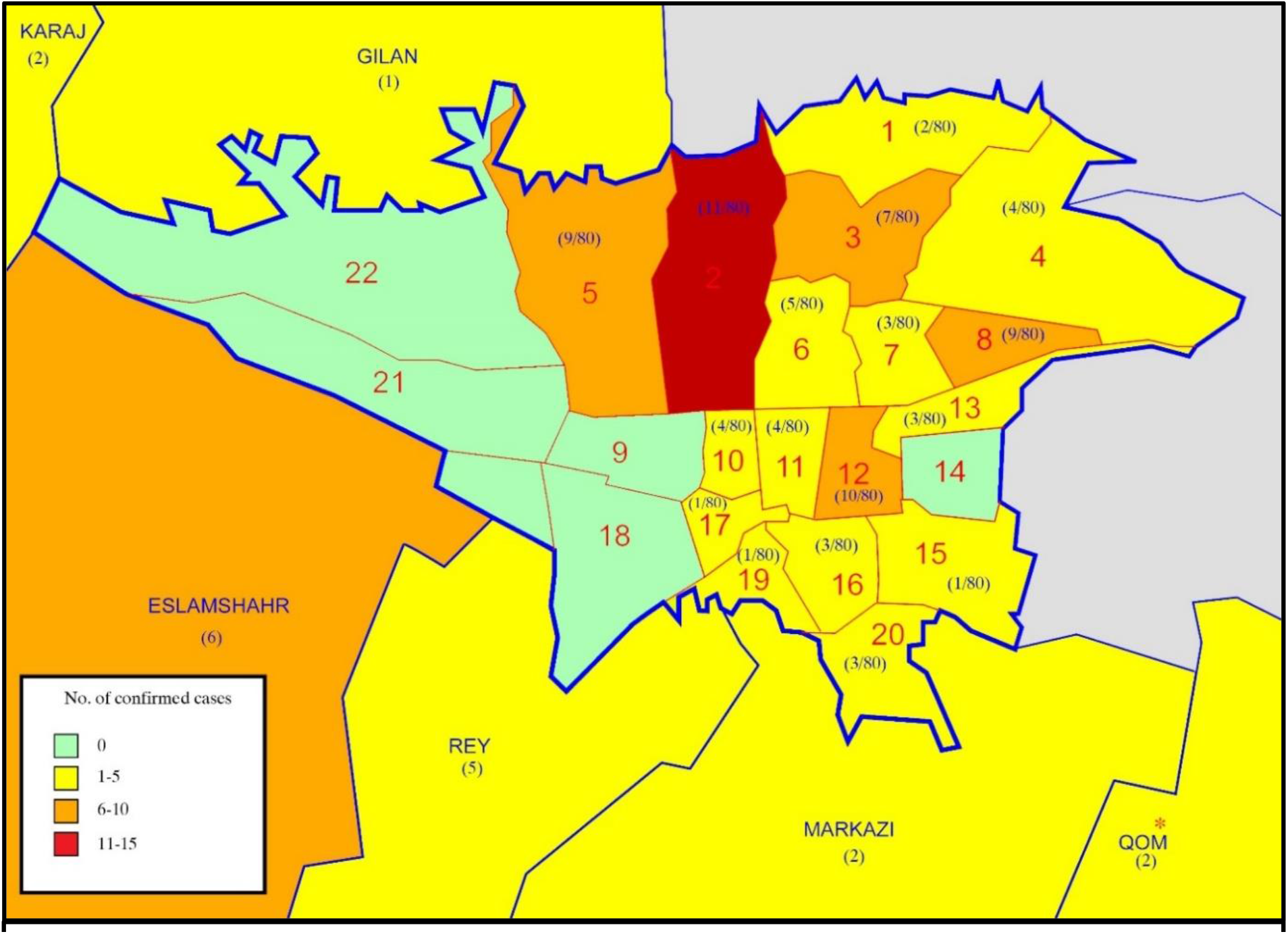
Disease Distribution Map in Tehran and Surrounding Areas/Cities. This map shows the distribution of all RT-PCR confirmed index patients in 22 districts of Tehran and surrounding areas. We did not have access to the address of two patients in the study. * Qom is marked as the epicenter of Covid-19 in Iran.

### Pre-hospitalization and demographic information

The median age of the patients was 58 years (range, 26-93). The majority of the patients (72.7%) were above 50 years of age. Critically ill patients were older than the non-critically ill group (100% vs. 67.9%; P= 0.005). Males constituted the majority of the patients (64.6%). The median of family members was 2 persons (IQR, 2-3) in a household. A total of 126 family members (55% female, 45% male) were identified to live in a household with index patients; 63% were above 50 years of age. According to job classification, 28 patients (28%) had low exposure risk occupations, 25 (25%) had high exposure risk occupations, and 5 of them (5%) were medical staff. Most potential exposures were contact with a suspected family member (22%) and contact with underage family members who had upper respiratory infection symptoms (8%). Nineteen patients (19%) who lived in Tehran had a recent history of domestic travel, and 3 (3%) had recent overseas travel. None of the patients recently traveled to or from China (table1).

### Clinical and paraclinical findings

The median incubation period was 7 days (IQR, 5-7). Fever was present in 45.2% of the patients on admission. The most common clinical symptoms were shortness of breath (74%), cough (68%), and myalgia (18%). Decrease level of consciousness was evident in 33% among critically ill patients, as compared with 0% among the non-critically ill group (P<0.001). Furthermore, respiratory rate was higher in critically ill patients compared with non-critically ill group (median of 25.5 vs. 19/minutes; P= 0.02). The presence of a coexisting disorder was higher in the critically ill group but was not statistically significant (73.3% vs. 60%, relative risk for the critically ill group, 0.59; 95% confidence interval [CI], 0.20-1.73; P= 0.25).

Laboratory tests on admission show that 74.2 % of the patients had lymphocytopenia, 92.3% had elevated C-reactive protein, 82.9% had elevated erythrocyte sedimentation rate, and 75% had elevated lactate dehydrogenase levels. The median level of white-cell count and median neutrophil count were statistically different in two groups of critically and non-critically ill patients. (P= 0.001 and P<0.001, respectively). Abnormal creatinine level percentage was higher in critical patients compared to non-critical ones (relative risk for the critically ill group, 4.53; 95% confidence interval [CI], 1.75-11.73, P=0.004).

In total, 55 CT scans were reviewed and scored by an expert radiologist. Non-parametric Friedman test shows different involvement in terms of lobar predominance. Right lower and left lower lobes were the most involved lobes followed by the right middle lobe, right upper lobe, and left upper lobe, respectively (P<0.001). Also, the test shows a difference in three cumulative scores. Median Lower lobes score was the highest score followed by median upper lobes score and median middle lobe score, respectively (P<0.001). Ground-glass opacity was the most common radiology finding (81.8%), followed by mixed pattern (ground-glass opacity + consolidation) and crazy paving appearance, which were found equally in the results (18.2%). Both groups (critically ill vs. non-critically ill) had similar CT scan findings.

### Treatment and clinical outcomes

All of the patients received oseltamivir as a recommended medication according to the national guideline. Other main administered medications included hydroxychloroquine (94%), lopinavir/ritonavir (60%), and ribavirin (12%) was administered in the patients. Intravenous antibiotics were also administered as shown in table 1. All patients received supplementary oxygen therapy based on patients’ conditions. Intravenous fluid therapy was given for routine maintenance, as mentioned by solution type and volume (table 1). In total, 19 patients were already taking losartan and angiotensin-converting-enzyme inhibitors (ACE inhibitors) due to hypertension, which continued during hospitalization course (16% losartan vs. 3% ACE inhibitors). Mechanical ventilation was used in 13% of the patients (2% non-invasive ventilation vs. 12% invasive ventilation).

**Table 1.** Demographic characteristics and paraclinical findings of hospitalized patients, Compared between critically ill patients and Non-critically ill patients.

| Variable | All Patients<br>(N=100) | Non-critically ill<br>(N=85) | Critically ill<br>(N=15) | P Value |
| --- | --- | --- | --- | --- |
| **Exposure history — no./total no. (%)** |  |  |  |  |
| Contact with suspected COVID-19 family member | 22/88(25) | 21/83(25.3) | 1/5(20) | 0.63 |
| Contact with medical staff in family member | 6/88(6.8) | 6/83(7.2) | 0/5(0) | 0.70 |
| contact with underage with upper respiratory infection symptoms | 8/88(9.1) | 8/83(9.6) | 0/5(0) | 0.61 |
| Contact with animals | 4/88(4.5) | 4/83(4.8) | 0/5(0) | 0.79 |
| **The use of preventing measures — no./total no. (%) <sup>a</sup>** |  |  |  |  |
| Used to wear medical masks | 5/88(5.7) | 5/83(6) | 0/5(0) | 0.74 |
| Used an alcohol-based hand rub | 9/88(10.2) | 9/83(10.8) | 0/5(0) | 0.58 |
| Used to wash hands regularly | 37/88(42) | 36/83(43.4) | 1/5(20) | 0.30 |
| Had personal knowledge about the disease symptom | 22/88(25) | 22/83(26.5) | 0/5(0) | 0.23 |
| **Travel history within 14 days before the onset of the symptoms — no./total no. (%)** |  |  |  |  |
| Domestic travel history | 19/88(21.6) | 19/83(22.9) | 0/5(0) | 0.29 |
| International travel history (except china) | 3/88(3.4) | 3/83(3.6) | 0/5(0) | 0.84 |
| Travel to china | 0/88(0) | 0/83(0) | 0/5(0) |  |
| **Social history — no./total no. (%)** |  |  |  |  |
| Smoker | 15/88(17) | 15/83(18.1) | 0/5(0) | 0.38 |
| Vaccination history | 8/88(9.1) | 8/83(9.6) | 0/5(0) | 0.61 |
| **Index patients Job classification — no./total no. (%) <sup>b</sup>** |  |  |  |  |
| Low exposure risk occupations | 28/58(48.3) | 27/57(47.4) | 1/1(100) |  |
| High exposure risk occupation | 25/58(43.1) | 25/57(43.9) | 0/1(0) |  |
| Medical staff | 5/58(8.6) | 5/57(8.8) | 0/1(0) |  |
| **Demographic information** |  |  |  |  |
| Age |  |  |  |  |
| median (IQR) — yr | 58(48-68) | 57(47-68) | 59(53-67) | 0.32 |
| Distribution — no./ total no. (%) |  |  |  |  |
| >50 — no. (%) | 72/99(72.7) | 57/84(67.9) | 15/15(100) | 0.005 |
| Male sex — no./ total no. (%) | 64/99(64.6) | 55/85(64.7) | 9/14(64.3) | 0.60 |
| Median hospitalization period (IQR) — days | 4(3-5) | 4(3-5) | 5(4-8) | 0.006 |
| Median incubation period (IQR) — days | 7(5-7) | 7(4-8) | 7(5-7) | 0.95 |
| **Vital signs on admission** |  |  |  |  |
| **Fever on admission — no. / total no. (%) <sup>c</sup>** |  |  |  |  |
| Median temperature (IQR) — °C | 37.5(37-38) | 37.5(37-38) | 38.2(37.1-38.8) | 0.12 |
| Distribution of temperature — no./total no. (%) |  |  |  |  |
| ≥37.8 °C | 42/93(45.2) | 34/81(42) | 8/12(66.7) | 0.10 |
| **Peripheral capillary oxygen saturation (SpO2) % — no./total no. (%)** |  |  |  |  |
| SpO2<93 % | 84/97(86.6) | 70/83(84.3) | 14/14(100) | 0.11 |
| **Median respiratory rate (IQR) — /minutes** | 19.5(18-22) | 19(18-21.25) | 25.5(18-28.5) | 0.02 |
| **Median heart rate (IQR) — /minutes** | 88(80-100) | 88(80-93.75) | 104(80.75-117.75) | 0.02 |
| **Blood pressure** |  |  |  |  |
| Median systolic blood pressure (IQR) — mm Hg | 110(100-130) | 110(100-130) | 110(104.5-135) | 0.81 |
| Median diastolic blood pressure (IQR) — mm Hg | 75(70-80) | 80(70-80) | 75(60-80) | 0.53 |
| **Clinical symptoms — no./total no. (%)** |  |  |  |  |
| Cough | 68/100(68) | 57/85(67.1) | 11/15(73.3) | 0.44 |
| Sputum production | 6/100(6) | 4/85(4.7) | 2/15(13.3) | 0.22 |
| Shortness of breath | 74/100(74) | 63/85(74.1) | 11/15(73.3) | 0.59 |
| Myalgia | 18/100(18) | 15/85(17.6) | 3/15(20) | 0.56 |
| Headache | 4/100(4) | 4/85(4.7) | 0/15(0) | 0.52 |
| Fatigue | 5/100(5) | 5/85(5.9) | 0/15(0) | 0.44 |
| Pleuritic chest pain | 11/100(11) | 7/85(8.2) | 4/15(26.7) | 0.06 |
| Rhinorrhea | 100/0(0) | 85/0(0) | 15/0(0) | -- |
| Sore throat | 4/100(4) | 2/85(2.4) | 2/15(13.3) | 0.11 |
| Nausea or vomiting | 1/100(1) | 0/85(0) | 1/15(6.7) | 0.15 |
| Diarrhea | 6/100(6) | 5/85(5.9) | 1/15(6.7) | 0.63 |
| Decrease level of consciousness | 5/100(5) | 0/85(0) | 5/15(33.3) | <0.001 |
| **Coexisting disorder — no./total no. (%)** |  |  |  |  |
| Any | 62/100(62) | 51/85(60) | 11/15(73.3) | 0.25 |
| Diabetes | 26/100(26) | 20/85(23.5) | 6/15(40) | 0.15 |
| Hypertension | 26/100(26) | 19/85(22.4) | 7/15(46.7) | 0.05 |
| Ischemic heart disease | 19/100(19) | 15/85(17.6) | 4/15(26.7) | 0.31 |
| Chronic obstructive pulmonary disease /asthma | 13/100(13) | 12/85(14.1) | 1/15(6.7) | 0.38 |
| Hypothyroidism | 6/100(6) | 6/85(7.1) | 0/15(0) | 0.37 |
| Others | 19/100(19) | 13/85(15.3) | 6/15(40) | 0.04 |
| **Laboratory findings** |  |  |  |  |
| **White-cell count** |  |  |  |  |
| Median (IQR) — per mm<sup>3</sup> | 6400<br>(4445-8525) | 5900<br>(4400-7775) | 12200<br>(6947.5-13525) | <0.001 |
| Distribution — no./total no. (%) |  |  |  |  |
| <4000 per mm<sup>3</sup> | 11/90(12.2) | 10/76(13.2) | 1/14(7.1) |  |
| 4000-10000 per mm<sup>3</sup> | 67/90(74.4) | 63/76(82.9) | 4/14(28.6) |  |
| >10000 per mm<sup>3</sup> | 16/90(17.8) | 7/76(9.2) | 9/14(64.3) |  |
| **Lymphocyte count<sup>d</sup>** |  |  |  |  |
| Median (IQR) — per mm<sup>3</sup> | 1100(849.5-1530) | 1100(848-1541) | 1248.5(918.6-1460.8) | 0.73 |
| Distribution — no./total no. (%) |  |  |  |  |
| <1500 per mm<sup>3</sup> | 66/89(74.2) | 54/75(72) | 12/14(85.7) | 0.24 |
| **Neutrophil count** |  |  |  |  |
| Median (IQR) — per mm<sup>3</sup> | 4510.2<br>(3244.8-6708) | 4237<br>(3201.3-6205) | 10505.5<br>(5225.4-12014.5) | <0.001 |
| Distribution — no./total no. (%) |  |  |  |  |
| >1800 per mm<sup>3</sup> | 7/90(7.8) | 6/76(7.9) | 1/14(7.1) |  |
| 1800-7800 per mm<sup>3</sup> | 67/90(74.4) | 63/76(82.9) | 4/14(28.6) |  |
| <7800 per mm<sup>3</sup> | 16/90(17.8) | 7/76(9.2) | 9/14(64.3) |  |
| **Platelet count<sup>e</sup>** |  |  |  |  |
| Median (IQR) — per mm<sup>3</sup> | 180000<br>(150000-214000) | 174000<br>(150000-213500) | 184000<br>(147250-268750) | 0.64 |
| <150000 per mm<sup>3</sup> | 20/87(23) | 16/73(21.9) | 4/14(28.6) | 0.41 |
| **Distribution of other findings — no./total no. (%)** |  |  |  |  |
| **Erythrocyte sedimentation rate<sup>f</sup>** |  |  |  |  |
| Median (IQR) — mm/hour | 43(32.5-60) | 40.5(28.5-59) | 50(42.5-77) | 0.22 |
| Elevated mm/hour | 34/41(82.9) | 30/37(81.1) | 4/4(100) | 0.46 |
| **C-reactive protein** |  |  |  |  |
| Median (IQR) — mg/liter | 36(20-54.8) | 36(20-56.6) | 43.25(22-52.3) | 0.76 |
| >6 mg/liter | 60/65(92.3) | 50/55(90.9) | 10/10(100) | 0.42 |
| **Lactate dehydrogenase** |  |  |  |  |
| Median (IQR) — U/liter | 584(461.3-736.3) | 581(467.5-715) | 1500(381-1531) | 0.27 |
| >480 U/liter | 30/40(75) | 28/37(75.7) | 2/3(66.7) | 0.60 |
| **Aspartate aminotransferase** |  |  |  |  |
| Median (IQR) — U/liter | 45(30-56.3) | 41.5(30-55.5) | 51.5(39-64.3) | 0.22 |
| >40 U/liter | 17/30(56.7) | 12/24(50) | 5/6(83.3) | 0.16 |
| **Alanine aminotransferase** |  |  |  |  |
| Median (IQR) — U/liter | 28(22-34.3) | 28(19.8-33.8) | 28(25-50.3) | 0.47 |
| >40 U/liter | 5/30(16.7) | 3/24(12.5) | 2/6(33.3) | 0.25 |
| **Alkaline phosphatase** |  |  |  |  |
| Median (IQR) — U/liter | 186(135.5-225.5) | 180(116.8-207.5) | 235(195.5-522) | 0.05 |
| >140 U/liter | 15/20(75) | 11/16(68.8) | 4/4(100) | 0.28 |
| **Creatinine kinase** |  |  |  |  |
| >170 U/liter | 4/4(100) | 1/1(100) | 3/3(100) | -- |
| **Creatinine** |  |  |  |  |
| Median (IQR) — µmol/liter | 106.1<br>(88.4-132.6) | 106.1(86.2-123.8) | 150.3<br>(101.7-221.1) | 0.01 |
| ≥133 µmol/liter | 17/83(20.5) | 10/70(14.3) | 7/13(53.8) | 0.004 |
| **Prothrombin time** |  |  |  |  |
| Median (IQR) — second | 13(13-14.9) | 13(13-13) | 14.8(13.4-17.3) | 0.05 |
| >13 second | 14/16(87.5) | 10/12(83.3) | 4/4(100) | 0.55 |
| **Partial thromboplastin time** |  |  |  |  |
| Median (IQR) — second | 32(29-38.5) | 32(29-35) | 36(28.8-41.8) | 0.70 |
| >39 second | 3/13(23.1) | 1/9(11.1) | 2/4(50) | 0.20 |
| **International normalized ratio** |  |  |  |  |
| >1.2 | 3/15(20) | 1/11(9.1) | 2/4(50) | 0.15 |
| **Blood gas — no./total no. (%)** |  |  |  |  |
| Metabolic acidosis | 2/28(7.1) | 2/24(8.3) | 0/4(0) |  |
| Respiratory acidosis | 0/28(0) | 0/24(0) | 0/4(0) |  |
| Metabolic alkalosis | 2/28(7.1) | 2/24(8.3) | 0/4(0) |  |
| Respiratory alkalosis | 3/28(10.7) | 2/24(8.3) | 1/4(25) |  |
| Metabolic acidosis and Respiratory acidosis | 5/28(17.9) | 3/24(12.5) | 2/4(50) |  |
| Metabolic acidosis and Respiratory alkalosis | 4/28(14.3) | 4/24(16.7) | 0/4(0) |  |
| Metabolic alkalosis and Respiratory acidosis | 6/28(21.4) | 6/24(25) | 0/4(0) |  |
| Metabolic alkalosis and Respiratory alkalosis | 6/28(21.4) | 5/24(20.8) | 1/4(25) |  |
| **Minerals** |  |  |  |  |
| Median sodium (IQR) — mmol/liter | 134(131.8-136) | 134(132-136) | 134(129.5-135.5) | 0.36 |
| Median potassium (IQR) — mmol/liter | 4.1(3.8-4.5) | 4.1(3.8-4.5) | 4.1(3.1-4.5) | 0.36 |
| **Radiologic findings <sup>g</sup>** |  |  |  |  |
| **Lobar predominance — no./total no. (%)** |  |  |  |  |
| Right upper lobe | 51/55(92.7) | 45/48(93.8) | 6/7(85.7) | 0.43 |
| Right middle lobe | 50/55(90.9) | 45/48(93.8) | 5/7(71.4) | 0.12 |
| Right lower lobe | 53/55(96.4) | 46/48(95.8) | 7/7(100) | 0.76 |
| Left upper lobe | 49/55(89.1) | 43/48(89.6) | 6/7(85.7) | 0.58 |
| Left lower lobe | 53/55(96.4) | 46/48(95.8) | 7/7(100) | 0.76 |
| **Scoring** |  |  |  |  |
| Lobar scores |  |  |  |  |
| Median right upper lobe score (IQR) | 1(1-2) | 1(1-2) | 2(1-3) | 0.12 |
| Median right middle lobe score (IQR) | 2(2-2) | 2(1-2) | 2(0-3) | 0.83 |
| Median right lower lobe score (IQR) | 2(2-3) | 2(2-3) | 2(1-3) | 0.96 |
| Median left upper lobe score (IQR) | 1(1-2) | 1(1-2) | 2(1-3) | 0.11 |
| Median left lower lobe score (IQR) | 2(2-3) | 2(2-3) | 2(1-3) | 0.96 |
| Cumulative scores |  |  |  |  |
| Median total score (IQR) | 8(7-11) | 8(7-11) | 9(6-15) | 0.51 |
| Median lower lobes score (IQR) | 4(4-6) | 4(4-6) | 4(2-6) | 0.96 |
| Median middle lobe score (IQR) | 2(1-2) | 2(1-2) | 2(0-3) | 0.83 |
| Median upper lobes score (IQR) | 2(2-4) | 2(2-4) | 4(2-6) | 0.12 |
| **Anatomic distribution — no./total no. (%)** |  |  |  |  |
| peripheral (subpleural) predominance | 55/55(100) | 48/48(100) | 7/7(100) |  |
| Central/perihilar predominance | 33/55(60) | 28/48(58.3) | 5/7(71.4) | 0.41 |
| Unilateral | 1/39(2.6) | 1/36(2.8) | 0/3(0) | 0.92 |
| Bilateral | 38/39(97.4) | 35/36(97.2) | 3/3(100) |  |
| **Attenuation** |  |  |  |  |
| Ground-glass opacity | 45/55(81.8) | 39/48(81.3) | 6/7(85.7) | 0.66 |
| Mixed<br>(ground-glass opacity and consolidation) | 10/55(18.2) | 9/48(18.8) | 1/7(14.3) |  |
| Crazy paving appearance | 10/55(18.2) | 7/48(14.6) | 3/7(42.9) | 0.10 |
| **Other signs** |  |  |  |  |
| Reticulation | 1/55(1.8) | 1/48(2.1) | 0/7(0) | 0.87 |
| Cavitation | 0/55(0) | 0/48(0) | 0/7(0) |  |
| Bronchiectasis | 0/55(0) | 0/48(0) | 0/7(0) |  |
| Pleural effusion | 4/55(7.3) | 2/48(4.2) | 2/7(28.6) | 0.07 |
| Lymphadenopathy | 2/55(3.6) | 2/48(4.2) | 0/7(0) | 0.76 |
| **Treatments** |  |  |  |  |
| Admission to intensive care unit— no. (%) | 12/100(12) | 0/85(0) | 12/15(80) | <0.001 |
| Mechanical ventilation — no. (%) | 13/100(14) | 0/85(0) | 13/15(86.7) | <0.001 |
| Non-invasive ventilation | 2/100(2) | 0/85(0) | 2/15(13.3) | 0.02 |
| Invasive ventilation | 12/100(12) | 0/85(0) | 12/15(80) | <0.001 |
| **Medications** |  |  |  |  |
| Oseltamivir — no./total no. (%) | 100/100(100) | 85/85(100) | 15/15(100) |  |
| Hydroxychloroquine — no./total no. (%) | 94/100(94) | 80/85(94.1) | 14/15(93.3) | 0.63 |
| Lopinavir / Ritonavir — no./total no. (%) | 60/100(60) | 47/85(55.3) | 13/15(86.7) | 0.02 |
| Ribavirin — no./total no. (%) | 12/100(12) | 3/85(3.5) | 9/15(60) | <0.001 |
| Systemic glucocorticoids — no./total no. (%) | 4/100(4) | 1/85(1.2) | 3/15(20) | 0.01 |
| Losartan — no./total no. (%) | 16/100(16) | 14/85(16.5) | 2/15(13.3) | 0.56 |
| ACE inhibitor — no./total no. (%) | 3/100(3) | 1/85(1.2) | 2/15(13.3) | 0.06 |
| Levofloxacin — no./total no. (%) | 52/100(52) | 43/85(50.6) | 9/15(60) | 0.35 |
| Vancomycin — no./total no. (%) | 32/100(32) | 23/85(27.1) | 9/15(60) | 0.02 |
| Azithromycin — no./total no. (%) | 21/100(21) | 19/85(22.4) | 2/15(13.3) | 0.34 |
| Ceftriaxone — no./total no. (%) | 23/100(23) | 20/85(23.5) | 3/15(20) | 0.53 |
| Piperacillin-tazobactam — no./total no. (%) | 6/100(6) | 5/85(5.9) | 1/15(6.7) | 0.63 |
| Meropenem — no./total no. (%) | 6/100(6) | 2/85(2.4) | 4/15(26.7) | 0.004 |
| Imipenem — no./total no. (%) | 5/100(5) | 4/85(4.7) | 1/15(6.7) | 0.56 |
| Ciprofloxacin — no./total no. (%) | 3/100(3) | 1/85(1.2) | 2/15(13.3) | 0.06 |
| **Intravenous fluid therapy** |  |  |  |  |
| Solution type — no./total no. |  |  |  |  |
| Dextrose 3.3% -sodium chloride 0.3% | 24/86(27.9) | 21/73(28.8) | 3/13(23.1) |  |
| Sodium lactate | 5/86(5.8) | 5/73(6.8) | 0/13(0) |  |
| Sodium chloride 0.9% | 5/86(5.8) | 3/73(4.1) | 2/13(15.4) |  |
| Sodium chloride 0.45% | 46/86(53.5) | 39/73(53.4) | 7/13(53.8) |  |
| Dextrose 5% - saline 0.9% | 6/86(7) | 5/73(6.8) | 1/13(7.7) |  |
| Median Solution amount (IQR) — cc/24hours | 1500<br>(1000-2000) | 1500<br>(1000-2000) | 1500<br>(1250-2000) | 0.12 |
| **Clinical outcome at hospitalization data cut off — no./total no. (%)** |  |  |  |  |
| Still hospitalized | 18/100(18) | 18/85(21.2) | 0/15(0) |  |
| Discharged from hospital | 70/100(70) | 65/85(76.5) | 5/15(33.3) |  |
| Death | 12/100(12) | 2/85(2.4) | 10/15(66.7) |  |

<sup>a</sup> Preventive measures consisted of wearing a medical facial mask when in contact with the public. 2. To use an alcohol-based hand rub 3. To wash hands regularly according to the world health organization (WHO) guideline. <sup>19</sup>
<sup>b</sup> The patient's occupation risk was classified into three groups. 1.Low exposure occupations that do not require close contact (at least within 6 feet) with the general public. 2.High exposure occupations that have frequent close contact (at least within 6 feet) with the general public. 3. Medical staff occupation was defined as a job in which people work in close proximity (at least within 6 feet) to patients known or suspected of COVID-19 infection.<sup>25</sup>
<sup>c</sup> Fever was defined as an axillary body temperature of 37.8 °C or above.
<sup>d</sup> Lymphocytopenia was defined as lymphocyte count less than 1500.
<sup>e</sup> Thrombocytopenia was defined as a platelet count of less than 150000.
<sup>f</sup> ESR normal range is dependent on age and sex of the patients and defined as follows: For male individuals 50> years of age, the normal range is below 15; for >50 and <85 years of age, the normal range is below 20; and for >85 years of age, the normal range is below 30. For female individuals 50> years of age, the normal range is below 20; for >50 and <85 years of age, the normal range is below 30; and for >85 years of age, the normal range is below 42. Any values above the normal limits were defined as elevated ESR in the table.
<sup>g</sup> Data regarding CT scan were missing for 45 patients due to the fact that they were performed at outside referring hospitals.

Hydroxychloroquine (OR=61.859; 95% CI for OR, 9.009-424.722) and the interaction of lopinavir/ritonavir*age*severity (OR=0.922; 95% CI for OR, 0.887-0.958) had a significant effect on the odds ratio. However, the interaction of azithromycin by hydroxychloroquine did not have a significant effect on the model (OR= 0.917; 95% CI for OR, 0.00-4.34*10^9^). Table 2 shows the first and the last step of the backward elimination in regression analysis. The value of Nagelkerke’s R^2^ for the final model was 0.840, and Cox and Snell’s R^2^ was 0.630, which both values showed the goodness of fit in our model.

**Table 2.** The Results of Logistic Regression Using a Backward Wald Elimination of Variables (Response: Outcome) ^a^

|  |  |  |  |  |  |  |
| --- | --- | --- | --- | --- | --- | --- |
|  | Regression coefficient (B) | Standard error (S.E.) | P value | Odds Ratio (OR) | 95% Confidence interval for OR |  |
|  |  |  |  |  | Lower | Upper |
| **Step 1** |  |  |  |  |  |  |
| age | -0.006 | 0.023 | 0.81 | 0.994 | 0.950 | 1.041 |
| hospitalization period | -0.005 | 0.259 | 0.98 | 0.995 | 0.599 | 1.651 |
| Hydroxychloroquine(1) | 5.138 | 2.944 | 0.08 | 170.3 | 0.531 | 5.46E+04 |
| Ribavirin(1) | -1.854 | 4.555 | 0.68 | 0.157 | 0.000 | 1180.949 |
| Lopinavir / Ritonavir(1) | 0.858 | 1.829 | 0.64 | 2.359 | 0.065 | 85.041 |
| Intravenous antibiotics(1) | -1.212 | 3.085 | 0.69 | 0.298 | 0.001 | 125.876 |
| Hydroxychloroquine(1) by age by severity(1) | 0.332 | 758.358 | 1.00 | 1.394 | 0.000 | NA |
| Lopinavir / Ritonavir(1) by age by severity(1) | -0.639 | 766.387 | 1.00 | 0.528 | 0.000 | NA |
| Ribavirin(1) by age by severity(1) | 0.236 | 110.643 | 1.00 | 1.266 | 0.000 | 1.91E+94 |
| Diabetes(1) | -2.310 | 1.750 | 0.19 | 0.099 | 0.003 | 3.063 |
| Hypertension(1) | 2.513 | 2.062 | 0.22 | 12.338 | 0.217 | 702.334 |
| Chronic obstructive pulmonary disease/asthma(1) | 34.177 | 10895.718 | 1.00 | 6.96E+14 | 0.000 | NA |
| Azithromycin(1) by Hydroxychloroquine(1) | -0.087 | 11.366 | 0.99 | 0.917 | 0.000 | 4.34E+09 |
| Age by Azithromycin(1) by Hydroxychloroquine(1) | 0.028 | 0.207 | 0.89 | 1.028 | 0.685 | 1.544 |
| **Step 13** |  |  |  |  |  |  |
| Hydroxychloroquine(1) | 4.125 | 0.983 | <0.001 | 61.859 | 9.009 | 424.722 |
| Lopinavir / Ritonavir(1) by age by severity(1) | -0.081 | 0.020 | <0.001 | 0.922 | 0.887 | 0.958 |

<sup>a</sup> Complete 13 steps of Logistic Regression is provided in the supplementary file (supplementary table 2).

Of the 185 patients admitted to the hospital during the study period, only 100 patients were eligible. Of these 100, 12 patients (12%) died, and 70 patients (70%) discharged at the date of data cut off.

The causes of death were as follows: five patients due to acute respiratory distress syndrome, 2 patients died of septic shock, 2 patients died due to cardiac arrhythmia, and 1 died of pneumothorax. The two remaining patients died of sudden cardiac arrest.

### Post-discharge follow-up

Seventy patients were followed within 14 days of discharge date. Thirty-six patients (51.4%) had observed 14 days of home quarantine post-discharge. Symptoms had relapsed in 40% of the patients. Shortness of breath (13%) and cough (13%) were the most common symptoms of relapse after discharge. Six of the patients (8.6%) were readmitted to the hospital, and 3 patients (4.3%) died post-discharge (Table 3).

**Table 3.** Post-discharge Follow-up.

| **Table 3. Post-discharge Follow-up.** |  |
| --- | --- |
| **Variable** | **Discharged Patients <sup>a</sup>**<br>**(N=70)** |
| Observing home quarantine after discharge — no./total no. (%) <sup>b</sup> | 36/70(51.4) |
| **Post-discharge symptom relapse — no./total no. (%)** |  |
| Any | 28/70(40) |
| Fever | 3/70(4.3) |
| Sore throat | 3/70(4.3) |
| Loss of appetite | 2/70(2.9) |
| Dizziness | 2/70(2.9) |
| Shortness of breath | 13/70(18.6) |
| Cough | 13/70(18.6) |
| Fatigue | 4/70(5.7) |
| Myalgia | 3/70(4.3) |
| Nausea or vomiting | 4/70(5.7) |
| **Post-discharge outcome — no./total no. (%)** |  |
| Hospital readmission | 6/70(8.6) |
| Death <sup>c</sup> | 3/70(4.3) |
| Recovery | 61/70(87.1) |

<sup>a</sup> Only discharged patients were eligible for the telephone survey (N=70).
<sup>b</sup> The patients were asked whether they completed 14 days of home quarantine after discharge.
<sup>c</sup> We could not determine the cause of death in patients who died post-discharged.

## Discussion

Our hospital was the first center to care for the new COVID-19 cases appearing in Tehran, Iran. We presented the first 100 cases of COVID-19 patients in Tehran. We identified the most common source of exposure, detailed clinical and paraclinical findings, the clinical outcome of common proposed antiviral therapies, and post-discharge follow-up.

The most important findings consisted of the positive effect of hydroxychloroquine and lopinavir/ritonavir on the disease outcome. Our findings are in concordance with previous studies, where hydroxychloroquine showed efficacy in disease outcome.^13,14^ Furthermore, *Cao et al*. concluded that lopinavir/ritonavir is not efficacious for COVID-19; however, the data was not assessed in relation to individual patient parameters.^15^ Our regression model identified age as a determinant in responsiveness to lopinavir/ritonavir, with efficacy being related to younger ages. Age has been identified as an important determinant in the mortality from COVID-19, but we show that younger age is also a determinant in the responsiveness to anti-viral therapy with lopinavir/ritonavir. We also used the model to determine the efficacy of a combined azithromycin/hydroxychloroquine regimen and found that the combination was not significant in clinical outcomes. This is contrary to current protocols and a previous study.^16^

The second most significant finding was symptom relapse in 40% of patients after discharge. The most common relapsed symptoms were cough (18.6%) and shortness of breath (18.6%). Six patients (8.6%) were readmitted to the hospital, and 3 patients (4.3%) died after discharge. This emphasizes the need for a close follow-up after symptom improvement. *Lan et al*. showed that certain patients could recover and test negative, only to test positive again.^17,18^ This phenomenon might underlie the symptom rebound in our patients and might indicate that patients are still a source of transmission after recovering from COVID-19.

The next significant finding in our study was a greater prevalence of COVID-19 in higher socioeconomic neighborhoods. We would have expected the lower socioeconomic segments in Tehran to be more important in transmission, but in our study, we found the contrary. This may be explained by the greater number of crowded areas like shopping malls and hospitals in affluent areas in comparison to the less affluent areas.

Furthermore, the majority of the patients did not follow WHO preventive measures; only 5% used medical masks, 9% used an alcohol-based hand rub, and 37% washed their hands regularly. ^19^ This emphasizes the importance of preventive measures.

Fever was present in less than half (45.2%) of the patients on admission, while the most common clinical symptoms were shortness of breath (74%) and cough (68%). Our data on fever is similar to *Guan et al*. who reported 43.8% fever on admission and differs from *Chen et al*. and *Wang et al*. who reported 83% and 98.6%, respectively.^6-8^ This might indicate that fever is not a specific finding in COVID-19. However, the cough has been a consistent prominent clinical symptom in COVID-19.

The severity of disease was directly related to patients age over 50 years, higher respiratory rate, and decreased level of consciousness. This is consistent with previous studies.^20,21^

Lymphocytopenia was a common laboratory finding. It may serve as a more specific marker at the beginning of this infection considering previous studies.^6-8^ However, it was absent in 25% of our study population.

Abnormal creatinine levels, higher white cell count, and higher neutrophil count were seen in our critically ill patients. This may be explained by direct renal involvement, or fluid imbalance secondary to the critically ill status of the patients.^22^ Increased WBC count in critically ill patients with the predominance of neutrophils can be a sign of secondary bacterial infection.

Chest CT scans analysis revealed higher involvement in both lower lung lobes compared with right middle and upper lung lobes. The most common finding was ground-glass opacity (81.8%).^23^ The presence of ground-glass opacity and bilateral lower lobe involvement is the most common radiographic findings of these patients, similar to *Xu Et al*., and can be used as a diagnostic factor for COVID-19. ^24^

## Limitations

First, we did not have access to review all CT scans since some were performed at outside referring hospitals. Second, the limited number of laboratory studies were due to the high patient load and limited resources. Third, many patients were excluded due to the lack of PCR kits at the onset of the epidemic in Tehran. Fourth, some patient medical records were not complete due to the emergency situation. Fifth, many of the patients were unable to remember initial exposure. Sixth, we could not determine the cause of death in patients who died post-discharged.

COVID-19 can present with a heterogeneous pattern of non-specific findings but affects older individuals more adversely. There is a high risk of disease relapse and necessitates close monitoring of discharged patients. The rush is on to find an effective therapy. The medical community is actively testing numerous repurposed and novel drugs.

## Data Availability

All data regarding the manuscript and supplementary files are available and will be provided as request.

## Acknowledgements

We appreciate all the hospital staff for their support and dedication to patients care, and all the patients who consented to their information being reported.

## References

1. Greenberg SB (2016) Update on Human Rhinovirus and Coronavirus Infections. Seminars in respiratory and critical care medicine 37 (4):555–571. doi:10.1055/s-0036-1584797

2. Ksiazek TG, Erdman D, Goldsmith CS, Zaki SR, Peret T, Emery S, Tong S, Urbani C, Comer JA, Lim W, Rollin PE, Dowell SF, Ling AE, Humphrey CD, Shieh WJ, Guarner J, Paddock CD, Rota P, Fields B, DeRisi J, Yang JY, Cox N, Hughes JM, LeDuc JW, Bellini WJ, Anderson LJ (2003) A novel coronavirus associated with severe acute respiratory syndrome. The New England journal of medicine 348 (20):1953–1966. doi:10.1056/NEJMoa030781

3. Arabi YM, Balkhy HH, Hayden FG, Bouchama A, Luke T, Baillie JK, Al-Omari A, Hajeer AH, Senga M, Denison MR, Nguyen-Van-Tam JS, Shindo N, Bermingham A, Chappell JD, Van Kerkhove MD, Fowler RA (2017) Middle East Respiratory Syndrome. The New England journal of medicine 376 (6):584–594. doi:10.1056/NEJMsr1408795

4. Zhuang Z, Zhao S, Lin Q, Cao P, Lou Y, Yang L, He D (2020) Preliminary estimation of the novel coronavirus disease (COVID-19) cases in Iran: A modelling analysis based on overseas cases and air travel data. Int J Infect Dis. doi:10.1016/j.ijid.2020.03.019

5. Tuite AR, Bogoch, II, Sherbo R, Watts A, Fisman D, Khan K (2020) Estimation of Coronavirus Disease 2019 (COVID-19) Burden and Potential for International Dissemination of Infection From Iran. Ann Intern Med. doi:10.7326/m20-0696

6. Guan W-j, Ni Z-y, Hu Y, Liang W-h, Ou C-q, He J-x, Liu L, Shan H, Lei C-l, Hui DSC D. B, Li L-j, Zeng G, Yuen K-Y, Chen R-c, Tang C-l, Wang T, Chen P-y, Xiang J, Li S-y, Wang J-l, Liang Z-j, Peng Y-x, Wei L, Liu Y, Hu Y-h, Peng P, Wang J-m, Liu J-y, Chen Z, Li G, Zheng Z-j, Qiu S-q, Luo J, Ye C-j, Zhu S-y, Zhong N-s (2020) Clinical Characteristics of Coronavirus Disease 2019 in China. New England Journal of Medicine. doi:10.1056/NEJMoa2002032

7. Wang D, Hu B, Hu C, Zhu F, Liu X, Zhang J, Wang B, Xiang H, Cheng Z, Xiong Y, Zhao Y, Li Y, Wang X, Peng Z (2020) Clinical Characteristics of 138 Hospitalized Patients With 2019 Novel Coronavirus-Infected Pneumonia in Wuhan, China. Jama. doi:10.1001/jama.2020.1585

8. Chen N, Zhou M, Dong X, Qu J, Gong F, Han Y, Qiu Y, Wang J, Liu Y, Wei Y, Xia J, Yu T, Zhang X, Zhang L (2020) Epidemiological and clinical characteristics of 99 cases of 2019 novel coronavirus pneumonia in Wuhan, China: a descriptive study. Lancet (London, England) 395 (10223):507–513. doi:10.1016/s0140-6736(20)30211-7

9. Lipsitch M, Swerdlow DL, Finelli L (2020) Defining the Epidemiology of Covid-19 — Studies Needed. 382 (13):1194–1196. doi:10.1056/NEJMp2002125

10. Health and Treatment Deputy of the Ministry of Health and Medical Education (2020). Guideline for the diagnosis and treatment of COVID-19 in outpatients and inpatients. http://dme.behdasht.gov.ir/uploads/Felo_Tashkish.pdf

11. Corman VM, Landt O, Kaiser M, Molenkamp R, Meijer A, Chu DKW, Bleicker T, Brünink S, Schneider J, Schmidt ML, Mulders DGJC, Haagmans BL, van der Veer B, van den Brink S, Wijsman L, Goderski G, Romette J-L, Ellis J, Zambon M, Peiris M, Goossens H, Reusken C, Koopmans MPG, Drosten C (2020) Detection of 2019 novel coronavirus (2019-nCoV) by real-time RT-PCR. Euro surveillance : bulletin Europeen sur les maladies transmissibles = European communicable disease bulletin 25 (3):2000045. doi:10.2807/1560-7917.ES.2020.25.3.2000045

12. The Ministry of Health and Medical Education (2020). Death of two new coronavirus patients in Qom. https://behdasht.gov.ir/%D8%A7%D8%AE%D8%A8%D8%A7%D8%B1/%D9%81%D9%88%D8%AA-%D8%AF%D9%88-%D8%A8%DB%8C%D9%85%D8%A7%D8%B1-%D9%85%D8%A8%D8%AA%D9%84%D8%A7-%D8%A8%D9%87-%DA%A9%D8%B1%D9%88%D9%86%D8%A7%D9%88%DB%8C%D8%B1%D9%88%D8%B3-%D8%AC%D8%AF%DB%8C%D8%AF-%D8%AF%D8%B1-%D9%82%D9%85

13. Gao J, Tian Z, Yang X (2020) Breakthrough: Chloroquine phosphate has shown apparent efficacy in treatment of COVID-19 associated pneumonia in clinical studies. Biosci Trends 14 (1):72–73. doi:10.5582/bst.2020.01047

14. Colson P, Rolain JM, Lagier JC, Brouqui P, Raoult D (2020) Chloroquine and hydroxychloroquine as available weapons to fight COVID-19. Int J Antimicrob Agents:105932. doi:10.1016/j.ijantimicag.2020.105932

15. Cao B, Wang Y, Wen D, Liu W, Wang J, Fan G, Ruan L, Song B, Cai Y, Wei M, Li X, Xia J, Chen N, Xiang J, Yu T, Bai T, Xie X, Zhang L, Li C, Yuan Y, Chen H, Li H, Huang H, Tu S, Gong F, Liu Y, Wei Y, Dong C, Zhou F, Gu X, Xu J, Liu Z, Zhang Y, Li H, Shang L, Wang K, Li K, Zhou X, Dong X, Qu Z, Lu S, Hu X, Ruan S, Luo S, Wu J, Peng L, Cheng F, Pan L, Zou J, Jia C, Wang J, Liu X, Wang S, Wu X, Ge Q, He J, Zhan H, Qiu F, Guo L, Huang C, Jaki T, Hayden FG, Horby PW, Zhang D, Wang C (2020) A Trial of Lopinavir–Ritonavir in Adults Hospitalized with Severe Covid-19. New England Journal of Medicine. doi:10.1056/NEJMoa2001282

16. Gautret P, Lagier JC, Parola P, Hoang VT, Meddeb L, Mailhe M, Doudier B, Courjon J, Giordanengo V, Vieira VE, Dupont HT, Honore S, Colson P, Chabriere E, La Scola B, Rolain JM, Brouqui P, Raoult D (2020) Hydroxychloroquine and azithromycin as a treatment of COVID- 19: results of an open-label non-randomized clinical trial. Int J Antimicrob Agents:105949. doi:10.1016/j.ijantimicag.2020.105949

17. Lan L, Xu D, Ye G, Xia C, Wang S, Li Y, Xu H (2020) Positive RT-PCR Test Results in Patients Recovered From COVID-19. Jama. doi:10.1001/jama.2020.2783

18. Zhang JF, Yan K, Ye HH, Lin J, Zheng JJ, Cai T (2020) SARS-CoV-2 turned positive in a discharged patient with COVID-19 arouses concern regarding the present standard for discharge. Int J Infect Dis. doi:10.1016/j.ijid.2020.03.007

19. World Health Organization (2020) Rational use of personal protective equipment for coronavirus disease 2019 (COVID-19): interim guidance. https://apps.who.int/iris/bitstream/handle/10665/331215/WHO-2019-nCov-IPCPPE_use-2020.1-eng.pdf

20. Mao L, Wang M, Chen S, He Q, Chang J, Hong C, Zhou Y, Wang D, Li Y, Jin H, Hu B (2020) Neurological Manifestations of Hospitalized Patients with COVID-19 in Wuhan, China: a retrospective case series study. medRxiv:2020.2002.2022.20026500. doi:10.1101/2020.02.22.20026500

21. Zhou F, Yu T, Du R, Fan G, Liu Y, Liu Z, Xiang J, Wang Y, Song B, Gu X, Guan L, Wei Y, Li H, Wu X, Xu J, Tu S, Zhang Y, Chen H, Cao B (2020) Clinical course and risk factors for mortality of adult inpatients with COVID-19 in Wuhan, China: a retrospective cohort study. Lancet (London, England) 395 (10229):1054–1062. doi:10.1016/s0140-6736(20)30566-3

22. Cheng Y, Luo R, Wang K, Zhang M, Wang Z, Dong L, Li J, Yao Y, Ge S, Xu G (2020) Kidney impairment is associated with in-hospital death of COVID-19 patients. medRxiv:2020.2002.2018.20023242. doi:10.1101/2020.02.18.20023242

23. Li K, Wu J, Wu F, Guo D, Chen L, Fang Z, Li C (2020) The Clinical and Chest CT Features Associated with Severe and Critical COVID-19 Pneumonia. Invest Radiol. doi:10.1097/rli.0000000000000672

24. Xu YH, Dong JH, An WM, Lv XY, Yin XP, Zhang JZ, Dong L, Ma X, Zhang HJ, Gao BL (2020) Clinical and computed tomographic imaging features of novel coronavirus pneumonia caused by SARS-CoV-2. J Infect 80 (4):394–400. doi:10.1016/j.jinf.2020.02.017

25. Occupational Safety and Health Administration (2020). Guidance for preparing workplaces for COVID-19. (Standard No. 3990-03). https://www.osha.gov/Publications/OSHA3990.pdf

